# Intersecting Vulnerabilities: Unraveling the Complex Relationship between Women, Climate Change, and Mental Health in Bangladeshi Communities

**DOI:** 10.1101/2023.06.06.23290721

**Authors:** Jean-Marc Goudet, Faria Binte Arif, Hasan Owais, Helal Uddin Ahmed, Valéry Ridde

**Affiliations:** Ceped, Université Paris Cité, Inserm, IRD, Paris, France; BRAC James P Grant School of Public Health, BRAC University, Dhaka, Bangladesh; National Institute of Mental Health and Hospital, Dhaka, Bangladesh; Institut de Santé et Développement, Université Cheikh Anta Diop, Dakar, Senegal

## Abstract

Climate change is one of the biggest challenges that humanity is facing in the 21st century, and its impact is being felt all over the world. It is well documented that the impacts of climate change have a significant effect on human health, including mental health. This paper focuses on the gender effect of climate impacts on mental health based on qualitative study conducted in two fragile communities in Bangladesh. This study was conducted using qualitative methods, including participant observation, in-depth interviews, and focus group discussions. The study was carried out in two fragile communities in Bangladesh, which were selected based on their vulnerability to climate change impacts. The participants were selected using snowball sampling. A total of 59 interviews and 3 Focus Group Discussions (FGDs) were conducted. Climate change impacts have a significant effect on mental health in both men and women. However, there are gender differences in the experience of climate change impacts on mental health. Women are more vulnerable to climate change impacts on mental health due to their gender roles and responsibilities. Responsible for taking care of their families, they have to face additional challenges due to climate change impacts, such as increased workload, water scarcity, and food insecurity, social insecurity as many of their husband migrates to the cities for jobs. Women also face social and cultural barriers, which exacerbates their vulnerability to climate change impacts on mental health. Men, on the other hand, face challenges related to their livelihoods and economic security due to climate change impacts. This study highlights the gender differences in the experience of climate change impacts on mental health in two fragile communities in Bangladesh. Socioeconomic and environmental determinants appear to be embedded and lead to psychological suffering in relation to social roles and gender norms. Interventions should be designed to address the specific needs and challenges faced by women in these communities. Policymakers should take a gender-sensitive approach to address the mental health impacts of climate change in these communities. This study contributes to the growing body of research on the gendered impacts of climate change with a trajectory approach and provides insights for future research in this area.

## Introduction

Since the end of the 2000s, climate change (CC) effects on mental health are described as a global emergency. Berry et al., described two main causal pathways between mental health outcomes and climate impacts [1]. Direct consequences occur after acute events and are described in terms of depression or psychological trauma [2]. Indirect consequences could be caused by the changes in ecosystems through different determinants such as economy, migration, or social structures [2]. For example, long-term changes such as chronic drought increased the distress of the rural population [3]. Additionally, the cumulative effects of multiple stressors associated with climate change can intensify existing mental health issues or lead to the emergence of new ones. Such stressors may include food and water insecurity[4–6], loss of livelihoods [7], displacement [8], and social networks [9]. Yet, a knowledge gap remains to characterize the chronic effects of slow-onset events on mental health issues [10]. Siegele defines the slow-onset events as phenomena with chronic changes (increasing temperature, sea-level rise, salinization, ocean acidification, loss of biodiversity, etc.) as opposed to acute events (storms, floods, acute drought, etc.) who are not permanent and occurred in a short time frame [11]. As the slow-onsets and their chronic consequences will keep rising, it is important to better understand the underlying mechanisms to provide adequate actions. Moreover, the slow onset events are disproportionally affecting the population living in the global South as they are more dependent on natural resources such as water and crops. While women are considered at higher risk to suffer from post-traumatic stress disorder after any natural disaster [12], how climate changes affect their mental health remain underexplored in global South. Recent result show how climate change, and specifically extreme weather events do affect the social structure. Women’s vulnerabilities are shaped by cultural and gender norms, as well as their lower socio-economic status in society, which manifests itself in their domestic role, decision-making within the household, and it is accentuated during climatic events [13] and in the context of a changing climate [14]. Indeed, these events prompt men to leave their communities in search of income, while women are trapped behind to shoulder caregiving responsibilities and additional duties [15]. But the underlying mechanisms of action are not well-explained. Moreover, there is limited data available from countries in the Global South, which are often the most vulnerable to the impacts of climate change.

Bangladesh is one of the countries most exposed to climate events (cyclones, tidal surges, floods, periods of drought) [16], and over the past decade, acute events have become increasingly frequent and intense due to climate change [17]. Simultaneously, the nation faces slow-onset events (acidification, increasing temperature, salinization, loss of biodiversity, etc.) that have led to environmental disruptions [18,19]. More than 60% of the population lives in rural areas and is significantly affected by slow-onset events [20]. The country is now confronting a dual crisis with the emergence of a mental health crisis. Recent studies highlight the substantial mental health burden in Bangladesh, with a 2019 average prevalence of mental disorders in the population reaching 16.8% [21]. A 2020 study published showed that 20 % of rural women were screened for a major depression disorder [22]. However, mental health infrastructure is limited, and very few psychiatrists are available in the country, with an estimated 260 psychiatrists for the 162 million people living in the area [23]. Moreover, research in the mental health field is underfunded and women’s mental health has been identified as one of the scientific priorities [24].

Our objective is to understand how mental health’s women are disproportionally affected by climate change in the context of vulnerable communities in the global South.

## Methodology

We conducted a qualitative research including participant observation, in-depth interviews, and focus group discussions [25].

This study is nested withing the Clim-HB project, a large public health research that focuses on health system resilience and climate change [26]. Our research took place in two vulnerable communities in Bangladesh, one in a rural area and the other in an urban area chosen due to their vulnerability and exposure to climate change.

In the rural community, the local population relies on agriculture and aquaculture (fish and shrimp culture) for their livelihoods. This community, located in the southwest region of the country, is subject to both slow-onset and acute climatic events. Moreover, 20% of the population experience a seasonal migration after the monsoon due to the lack of employment. The second setting is a slum in the capital city. The investigation primarily focused on the poorest households in the slum, who live in stilt houses built over a canal. This setting is particularly exposed to pollution and high temperatures. These field sites are part of a larger public health research project within the framework of the Clim HB project [26]. Both populations share a pattern of migration, as a large portion of the families encountered in the slum originated from coastal regions of Bangladesh and migrated for economic reasons, and sometimes environmental ones. Coastal regions are subject to significant environmental transformations as well as a scarcity of jobs, which is also related to the urbanization of the country [18,20].

The communities were selected using purposive sampling, and the participants were selected using snowball sampling. To reflect the seasonal variations that could influence mental health issues, data were collected during two period covering three months in total: in March and April during the hot season and in October 2022 after the monsoon. In each site, approximately fifteen households were followed throughout both data collection periods. We conducted interviews with various household members to triangulate the data, which helped improve the reliability and quality of the information gathered. This method proved to be essential in establishing the research relationship, allowing us to reconstruct social trajectories and assess the impact of specific environmental events on women’s mental health. Table 1 shows that the study comprised a total of 55 interviews and 6 focus group discussion (FGD). Only relevant data for the research question were retained for the analysis.

**Table 1.**
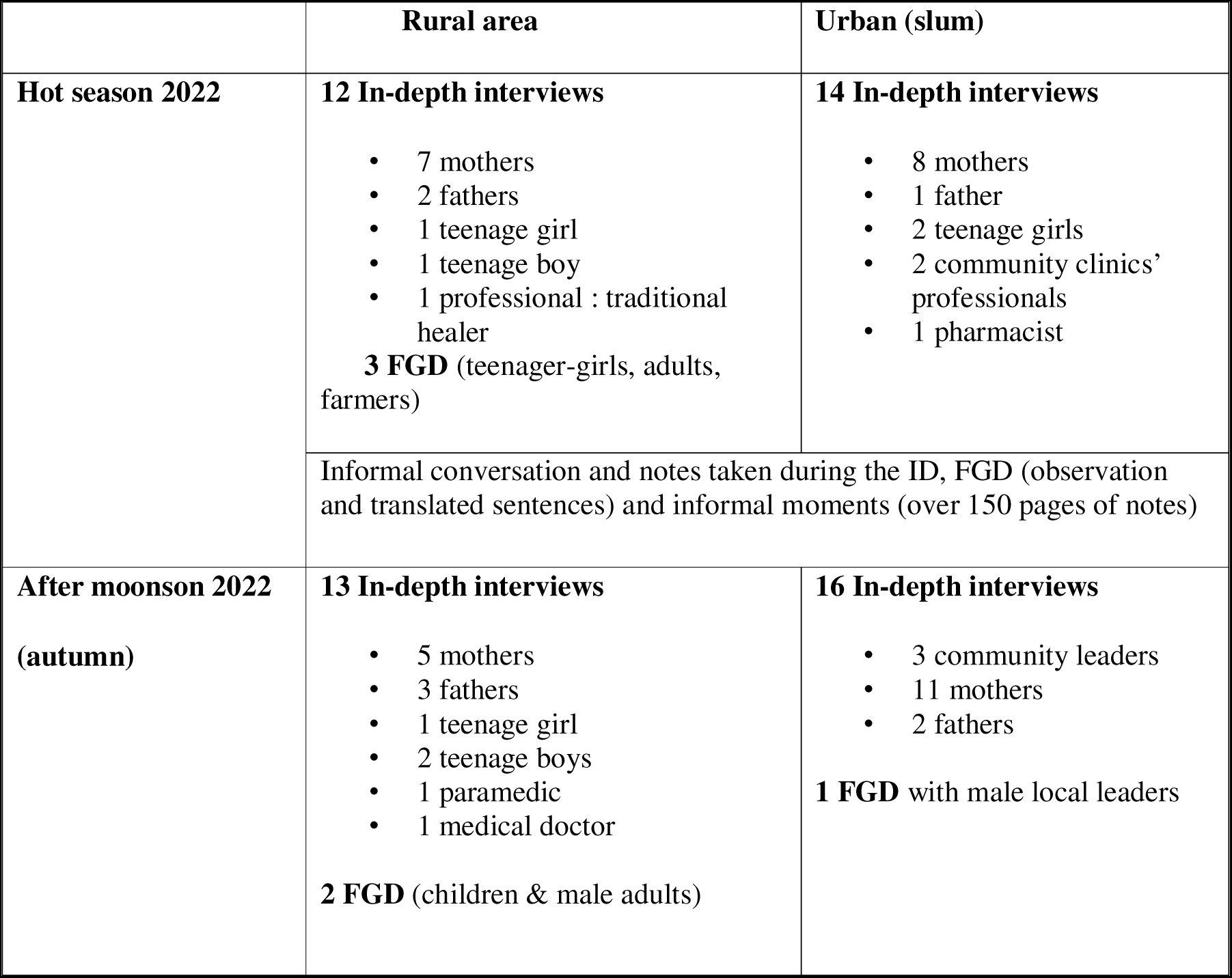
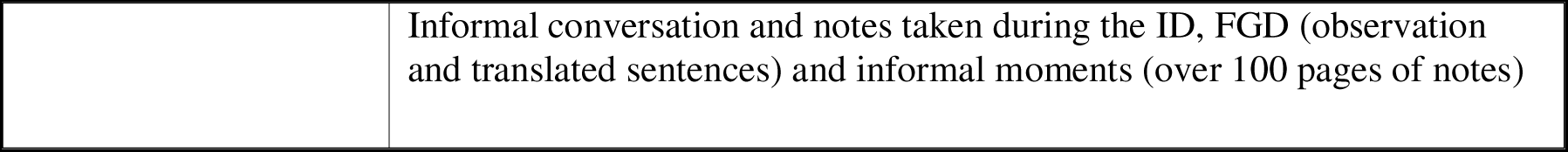
Number of interviews and FGDs by site and season of study, Bangladesh, 2022.

|  | Rural area | Urban (slum) |
| --- | --- | --- |
| Hot season 2022 | **12 In-depth interviews**<br>• 7 mothers<br>• 2 fathers<br>• 1 teenage girl<br>• 1 teenage boy<br>• 1 professional : traditional healer<br>**3 FGD** (teenager-girls, adults, farmers) | **14 In-depth interviews**<br>• 8 mothers<br>• 1 father<br>• 2 teenage girls<br>• 2 community clinics' professionals<br>• 1 pharmacist |
|  | Informal conversation and notes taken during the ID, FGD (observation and translated sentences) and informal moments (over 150 pages of notes) |  |
| After moonson 2022<br>(autumn) | **13 In-depth interviews**<br>• 5 mothers<br>• 3 fathers<br>• 1 teenage girl<br>• 2 teenage boys<br>• 1 paramedic<br>• 1 medical doctor<br>**2 FGD** (children & male adults) | **16 In-depth interviews**<br>• 3 community leaders<br>• 11 mothers<br>• 2 fathers<br>**1 FGD** with male local leaders |

|  |  |
| --- | --- |
|  | Informal conversation and notes taken during the ID, FGD (observation and translated sentences) and informal moments (over 100 pages of notes) |

The majority of the interviews were conducted in Bengali with the presence of a Bengali-English translator. During the second fieldwork, the researchers returned to the same individuals to conduct a follow-up and refine the data obtained from the first fieldwork. In each site, around fifteen families were interviewed. When possible, researchers conducted interviews with different household members, to triangulate and gain a comprehensive understanding of the family’s situation.

A thematic analysis was then performed based on the social determinants framework from WHO and Lund et al. [27,28]. To understand how power relations among gender are constituted in a changing climate, it was combined to specific dimension (access to resources / division of labour within and beyond the household) from the gender framework [29]. The authors had access to non-anonymized individual participant data, but it is important to note that the data presented in this study have undergone a rigorous anonymization process and have been maintained with strict confidentiality.

## Ethics declarations

All participants provided signed consent to participate. Written consent was obtained from both the parents and the individuals themselves for participants under the age of 18, ensuring that both parties were informed and gave their consent to take part of this study. The study protocol was reviewed and approved by the ethics committee from the Institutional Review Board (IRB) of the BRAC James P Grant School of Public Health, BRAC University (ref: IRBLJ19 November’20–050) in Bangladesh.

## Results

Two main pathways of mental health outcomes impacted by climate change through gender have been identified in these vulnerable communities. These pathways are interconnected, depending on the social and environmental situation, and they often manifest as cumulative effects that shape women’s trajectories.

### The socioeconomic stressor as an intermediate factor

This first pathway contributes to exacerbating pre-existing gender inequalities arising from economic determinants. It leads to anxiety and depressive-like disorders by amplifying individual vulnerabilities through the impact on livelihood.

### Impacts on mother responsibilities

Climate change directly impacts the livelihoods of the most vulnerable populations. Due to drought, peoples from rural setting are experiencing a decline in agricultural production and shrimp yield. The immediate effect on market prices leads to food insecurity that challenge mothers’ responsibilities. It results in increased anxiety for them and potential adverse effects on the physical health of their children. This affects all families in the studied village, including those with a higher socio-economic status.

> *I: There are some problems. Suppose we can now buy 1.5 kg when we could buy 2kg in the past. It has become like this due to the price hike.*
>
> *R: Any problems to feed for the children*
>
> *I: Even if we are starving, we have to feed them. We have to meet their need. (mother 1)*

This suffering is amplified in households that are already in economic difficulty with debt.

> *I: You explained this year it did not rain much and it rained heavily for only 2 or 3 days. Do you think you have faced any problems with the lack of rainfall?*
>
> *R: We have faced some problems with the lack of rainfall such as we had to irrigate our land manually, and we have faced problems regarding jute retting. (…) We also had problems with the money lender. So, we had to take money from our relatives as well. It was very stressful (mother 2)*

Those that are in-debt experience the situation as one of the most difficult in their lives. The situation seems worse for them due to a reduced ability to implement coping strategies. Indeed, the mother 1, whose husband belongs to a higher social class and has attempted to engage in local politics, holds significant social and symbolic capital in the village. The family has better economic resources and greater diversification. They own a pond with fish, a small vegetable garden, and a chicken coop. In the opposite, mother 2 and her husband have a small land and a little grocery store that experienced the COVID crisis, and they both come from impoverished families. Furthermore, mother 2 has two young children, one of whom she is breastfeeding. When asked about her sleep quality, she expresses difficulties related to her child’s crying, who is not adequately nourished.

> *I: The child annoys me a lot. I can’t breastfeed her properly (…)*
>
> *R: No, I could not sleep previously because of the financial crisis and now I cannot sleep properly because my child cries at night.*
>
> *I: Do you wake up a lot during the night*
>
> *R: He wakes up 2 or 3 times at night.(…) He does not like to eat rice. He always asks for good food like fish, and eggs. (mother 3)*

Thus, the effects of climate change tend to follow the socio-economic gradient and the preexistent social inequities. For mother 2, who belongs to a lower class and is heavily in debt, food insecurity leads to disruptions in breastfeeding and undermines her mental health as a caregiver. On the other hand, for mother 1, the psychological distress is less severe due to her financial capacity to cope with the situation. Due to hampering in the breastfeeding leads the nutrients lacking in her child also. The impacts are continuing in the cyclic pattern.

In each instance, these situations were described by the women who bear the responsibility of caring for and feeding the children, while the interviewed men explained their strategies for obtaining income without ever mentioning the mothers’ and children’s circumstances. This indicates the overlooking tendencies of the women’s burden also be discussable.

Similarly, their economic dependence on their husbands sometimes prevents them from leaving their marriage when they consider them unfit to fulfill their roles.

> *R: But after marriage, my husband once left his job and started pulling a rickshaw. He used to pull the rickshaw one day and sit at home for three days without earning any money. At that time, I got a feeling of leaving this family and staying separate from my husband. But I couldn’t do that because of my children. I was worried for them. (mother3)*

Income-generating activities in rural areas are not very popular. Due to their limited social capital and reduced agency, women often cannot leave their husbands to seek economic security for their children unless they can receive support from other family members, such as their own parents.

### Coping strategies on the household rely on the daughters’ & mothers’ sacrifices

Coping strategies for the economic situations are dependent on social roles within families and based on a division of labor. Both are exacerbated in a changing climate. Mothers, who are often housewives to raise and educate the children, bear the responsibility of caregiving and thus have fewer options for economic survival strategies. This specifically impacts their mental health.

One of the economic survival strategies presented by a mother from the slum involved not bringing all her children with her. She took the youngest child who was breastfeeding and needed her, while she left her eldest daughter with her mother-in-law. She tries to visit her daughter every three months but describes this situation as very painful for her.

> *I: How is your sleep now? Are you tensed about something?*
>
> *R: I am bound to be tensed about my son or my daughter. I can’t do any work.*
>
> *I: If you could tell us what are your thought about your children?*
>
> *R: The mother has to suffer the pain of the son. Will anyone else suffer?*
>
> *I: Can you sleep properly or you wake up from the sleep early?*
>
> *R: I do wake up suddenly. I think what I did to deserve this. (mother 4)*

She didn’t want to move to the slum but had to as her husband has job there. They are both from the coastal area where it became hard to survive. Economic migration was thought as a survival strategy decided by the husband.

Furthermore, this economic migration desired by her husband isolates her. Being far from her mother and sister, makes it even more difficult for her to cope with the situation. Besides, her inability to work as she has to take care of her son reinforce her feeling of isolation.

> *R: I have to think about the rent, food and everything. But in village, i can share everything with my mother and my sister, which is very much better than this. (mother 4)*

Additionally, she explains that she had to accept being married to financially relieve her parents and could not pursue her studies.

> *R: I have five siblings and I am the eldest among the sisters. I studied till class 6 and then my parents married me off to reduce the burden (…). (mother 4)*

Economic strategies for these poor rural families are mainly decided by men so they can provide for their families, even though this becomes increasingly complicated during times of economic and environmental crisis.

In some cases, environmental events shape the economic strategies of the families.

Another respondent explained, that after an environmental event, her father decided to married her.

> *R: When I was a child, I observed a lot of cyclones and storms.*
>
> *I: Did your house suffer damage during those cyclones?*
>
> *R: We had 2 houses, one of the fell apart because of the Cyclone. It happened before the river erosion. And we lost everything including houses in the river erosion.*
>
> *I: When did this river erosion happen?*
>
> *R: It happened around the mid-2000’s. Before my marriage.*
>
> *I: Did you got married for this reason or there were other problems in your family?*
>
> *R: Before the river erosion, my family had the land and we were financially strong but after the incident we got poor. And since we were so many siblings my father took the decision about my marriage.*
>
> *I: What type of land were those?*
>
> *R: They were agricultural land and we used to grow rice and other crops on them.*
>
> *I: Can you tell me the size of those lands?*
>
> *R: It was a good amount of land. The whole area was gone by river erosion. (mother 5)*

In this case, the environmental event leads to a descendant social mobility of the family. Now living in the slum, this mother face economic constrained that generate anxiety. She is concerned about her children education and a broader vision of their future.

> *R: It will be much more reliable for us to survive if my husband gets a new job. We pay 4,000 takas as rent for one room and, my husband and my children live with me in that one room.*
>
> *I: Can you tell me about your food expenses?*
>
> *R: Every month it costs about 10,000 to 12,000 taka for our food and children’s education combined. The house rent is separate. (…)*
>
> *R: I’m having some stress about the future of my children and, also since my husband has no job, I’m concerned about how we will finance the upcoming days. I’m having headaches because of this stress. (mother 5)*

### Gender violence

Although these situations have been documented outside the context of climate crises [30], they appear to be more intense and recurrent when crises affect the household’s livelihood and reduce the opportunities for men to generate sufficient income to meet the family’s needs.

For example, the previous woman who was married after a river erosion and came in the slum faced domestic violence because of the unemployed situation of her husband and his addiction.

> *R: I feel stressed now, but the reason is different. My husband has no work and he is addicted, because of that we have conflicts in the house and also many other domestic problems, that’s why I have stress and tension all the time. (…) As he used to spend the money on drugs and I tried to convince him, for that reason he used to beat me. (mother 5)*

Like other respondents, she is particularly isolated, which increases her stress. Her mother-in-law is her close neighbor, support her son, and would report to him any of her complaints. This exacerbates the situation and further contributes to the respondent’s stress and feelings of isolation.

> *R: I’m trying to convince my mother-in-law as well, but every time they say that he will quit his addiction soon but it’s not happening and these types of conversations are making conflicts between my husband and his mother and also me. because I’m informing my mother-in-law about my husband’s addiction and somehow, he is also getting to know that I informed his mother. That’s why there are conflicts between us also. (mother 5)*

Several women explained to us that when they discuss household money management with their husbands, particularly in situations of debt, the husbands reprimand them and may physically abuse them.

> *R: I used to have arguments with him, regarding the bazaar he used to take money from people, and if I ask questions about it then he was beating me.*
>
> *I: He is an alcoholic?*
>
> *R: No. He used to gossip with his friends and spent money, this is his habit, but our house condition is not good. That’s the reason*
>
> *I: When you used to say something that time he used to beat?*
>
> *R: I said that, we have children, and they can’t live without eating, he didn’t do the bazaar properly.*
>
> *I: He used to take loans?*
>
> *R: Yes.*
>
> *I: He used to take loans from people but he didn’t go to work, if I ask anything then he used to beat me. (mother 6)*

In the rural area, a family facing over-indebtedness also experienced the negative effects of climate change on their mental health. The woman explained that their financial situation was already poor before, but it worsened this year due to a lack of rain. The combination of financial stress and the impacts of climate change can create a challenging situation for families, particularly for women, mothers, and widows who bear the responsibility of caregiving.

> *R: We also had problems with the money lender. So, we had to take money from our relatives as well. It was very stressful.*
>
> *I: Do you sleep well at night?*
>
> *R: No, I could not sleep previously because of the financial crisis and now I cannot sleep properly because my child cries at night. (…)*
>
> *I: How is your relationship with your husband?*
>
> *R: The relationship is good but we get into fights for financial matters. People come over to ask for money and get into fights over it.*
>
> *I: Does he physically abuse you along with verbal abuse? R: He hits me at times.*
>
> *Hasan: When was the last time he hit you?*
>
> *R: It starts with an argument. I try to stay patient. People tend to pass judgment as I live in my father’s house. People will talk, that is why I need to remain patient. (mother 7)*

This inescapable situation, which is often the result of norms and social pressures placed on women regarding the importance of religious marriage, can significantly impact their mental health and well-being. Women may feel trapped in their circumstances, unable to leave their marriages or seek help due to the expectations and obligations associated with their roles as wives, mothers, and caregivers.

> *R: Because my elder brother is very angry, if he listened to that, my husband is shouting at me or if he is misbehaving with me, then he will beat him. That’s why I don’t.*
>
> *I: He doesn’t know that he used to beat you?*
>
> *R: Yes, he knows, he tried to get him arrested several times.*
>
> *I: Okay.*
>
> *R: But he couldn’t do it because of me. I said how many times people get married, once so let it be, let me stay with one person. (mother 7)*

#### Lack of coping strategy

The isolation experienced by many mothers and young girls in reinforced by the social norms that disregard discussions about psychological issue. This is mechanism that is both self-controlled and encouraged by men due to the significance of reputation within these communities. Reputation is not only important for marrying off their children but also for matters of social dignity.

> *R: I always think if I’m able to do this then, it will be better but I don’t have money, so how I’ll do it? My children wanted to live in a good place and to have good food, but I am unable to do anything. No one will help me, I didn’t share with anyone, my son asked me not to share because people will judge us, I am sharing with you guys as you’re listening otherwise, I don’t share with anyone, my son is different, he doesn’t share his sadness with anyone. He has grown up. So, he doesn’t like this. (mother 8)*

### Climate events as a triggering factor

The second mechanism highlighted by our data concerns the triggering effect of climate events, that affects young girls and mothers.

We had already encountered this issue with environmental event during our fieldwork in April 2022 following the COVID-19 pandemic. Some women had mentioned having suicidal thoughts during that period due to their perceived failure in fulfilling their caregiving responsibilities for their children.

> *R: During the covid situation, I and my two children were staying with my husband. Whenever our children asked for food, we were not able to provide them with food. That time I had a feeling of committing suicide.*
>
> *I: Have you ever tried to commit suicide?*
>
> *R: Yes, I tried to commit suicide once in the middle of the Corona situation. But with the poison bottle in my hand, I couldn’t commit suicide anymore when I thought about my children because I was worried about who’ll look after my children after I will be gone. (mother 9)*

### Hot weather and children behavior

During our first field in the hot season, indoor temperatures in the tin houses of the slum were very high and significantly higher than outdoor temperatures. Several women reported children’s sleep problems resulting into behavioral issues during the day. They explained that they could not control their children’s behavior, which led them into a negative spiral of increasing anxiety and stress, due to sleep deprivation, compounded by the children’s difficult-to-regulate behavior.

### Environmental & specific setting

The specificities of climate events are important to consider for the policy maker and in order to help the most vulnerable people. Usually the main characteristics retained are the duration, frequency, intensity of an event, geographical location such as coastal areas [31].

During our study, the cyclone Sitrang occurred and was considered by local authorities as having caused much damage due to its low intensity. However, the topography of the part of the slum involved in our study, whose houses are located on a canal, were highly affected. For the respondent, this cyclone that occurred in October 2022 was much more difficult than one of the last cyclones recognized as having caused the most damage in Bangladesh (Amphan in 2020), even if she was at this time living in the coastal area. Indeed, here she did not receive any assistance or rescue and was trapped in her room, although she took refuges during Amphan.

> *R: I think this cyclone was more dangerous. I stayed here the whole night. Yes, it was more dangerous for me (…) there we suffered from damage, one of our big trees was fallen into our house, our house was broken because of that, later we were unable to fix it, the government helped us, and send some dry food. (mother 10)*

The inability to access shelters for these impoverished families has made them more vulnerable, even though they have migrated from coastal areas that have been hit by multiple cyclones, but where a refuge had been accessible.

### Housing vulnerability in the face of climate events

This importance of housing was highlighted during the Cyclone Sitrang which occurred in October 2022 and caused significant damage to families living in the slum. Some of our respondents described the alarming situation for living in the tin houses located on the part of the slum built on stilts above a canal. During the cyclone, the rainwater arrived both from clouds and canal, whose water level began to rise suddenly in the evening. While the majority of family took refuge in a public building, this one found trapped in her room.

> *R: During the cyclone, our house got flooded. The water level was very high because our house was one of the low-level houses in this area. We lost many small household things such as dishes and other belongings during that flood. The water level rose and flooded our house at 11 pm night. The rain started early in the morning. We were sleeping in the house when the water got into our house. We didn’t expect that the water level would rise that much because previously when it rained very little amount of water used got into our house but that night the water level was very high. (mother 11).*

The situation was catastrophic as the family faced the prospect of death. While the mother recounts the extreme tension, her children were traumatized, and her daughter remains in shock several days after the event.

> *R: When we were standing in bed, I was very tense. We were afraid that if the rain continued, the water will increase, and we will be stuck here and die. One person came during that time and he was asking if someone was stuck here in these houses. We shouted yes and said that we were there. But I don’t know whether he listened to us or not because he didn’t respond. Also, there were so many mosquitoes biting us continuously, so we also asked that guy if he could bring us a mosquito coil. But he didn’t respond. And no one came to rescue us later on.*
>
> *I: How were the babies doing during that time?*
>
> *R: We removed the sewing machine from the table and placed the table on the bed. Then we kept our children on top of that table. They sat there the whole night. My daughter cried the whole night because she was very afraid of seeing the water. She is still sick. My son also got sick and was afraid, so I took him to maulana (the religious leader of the community), and now he’s feeling good. But my daughter is still afraid and sick. (mother 11)*

Housing is one of the most important factors in resilience or vulnerability to climate events (heat waves, floods or cyclones). In both rural and urban sites, the houses that were less exposed to flooding and more resilient to extreme weather conditions offered greater security and protected mental health outcomes. Here too, gender plays a significant role in the inheritance that is passed down to children, reflecting an inequality in the distribution of material resources. The previous respondent, who is abused by her husband, lives in a tin house, while her brother, who wants to help her, resides in their parents’ hard-building house. She is thus doubly vulnerable as she does not live in durable housing and she does not move because her husband is not a decent enough person for him to live with his family.

> *I: Why don’t you stay there?*
>
> *R: I don’t stay there as my husband is not good. Maybe he can say a bad word, they can hear that. It’s not good to stay with the relative (mother 11)*

### A single mother with mental health preconditions

One of the most pronounced vulnerabilities to climate events is being a single mother raising her children. One of our respondents describes the alarming situation she experienced when she was caught off guard by the rising waters. Her tin-roofed room is located on a part of the slum built on stilts above a canal. The rainwater arrived both from the clouds and from the canal, whose water level began to rise suddenly in the evening.

> *R: Everyone went there in the shelter, in our area, there was so much water, only me, my sister and my child were here, my sister told me that: “no one is here, do you want to die?”. Then somehow, I catch her and I started swimming, it was not the condition of walking. I started swimming, carrying my child on my shoulder, then my sister even got fallen due to the water pressure. So, I came back to this room. (mother 12)*

She didn’t go in a shelter as others because she perceived it a risk as a single mother without no other social support. Her brother also stayed in a tin house during the event.

> *R: We were only sitting on the bed, but we were unable to sleep.*
>
> *I: Only you and your son?*
>
> *R: Yes. Many people went from here, but I didn’t go as I don’t have any place to go, where I’ll go.*
>
> *I: During the night water level was increased, you did not do anything?*
>
> *R: Where I will go along with a child, so I didn’t go outside, I stayed here. I didn’t feel good to go any other place with my children, and it was hard for me to go anywhere to take the shelter as a single mother, so that is why I didn’t take the risk, I stay here on the bed only. (mother 12)*

She was also afraid by the current situation of water in the road and her inability to protect her son from the water.

> *I: As everyone leaves here, no one asks you to go with them?*
>
> *R: Yes, everyone was telling me to go from here, I said no, I’ll not go, I had the courage, and I said that, if my death is written here then I’ll die here along with my son, I’ll not go anywhere.*
>
> *I: Why you did not go anywhere?*
>
> *R: I didn’t go, because I didn’t feel good, I had the courage in my mind that nothing will happen, if my death is written here then I’ll die here, if I go there, I can also die there. I had the courage in my mind. (mother 12)*

Finally, she left her son with her uncle who stayed as the responsible person of the tong houses. Because her son was crying too much in her arms, as she was too stressed by the situation. Consequently, she stayed alone in the room, feeling very anxious, as she constantly thought about her son and found solers in her prayers.

> *R: I am million times thankful to Almighty Allah. Later on, the next day, the water level decreased and the rain almost stopped. So, as it was not raining, so I was less tensed. (…) I was so afraid when the water level was increasing, and I was thinking maybe it’s going to be the end, but I performed special prayers, and I prayed to Almighty Allah, to save us and also, I was also remembering my Almighty Allah’s name again and again, and then I saw in the morning the water level was decreasing. Around 3-4 AM it started less raining, at that time I was like the water level will not increase, and at that time I was having less stress. (mother 12)*

Her initial decision to stay alone in the tin house with her son, awaiting death, is linked to her pre-existing mental condition. This woman has experienced several depressive episodes, including one a few months before the cyclone, which left her bedridden for a month, preventing her from working and taking care of her son, who stayed with his uncle. Her psychological difficulties began during childhood, without her recalling the reason, and then following her marriage and upon discovering that she was her husband’s second wife. A few years later, her parents passed away, and her husband died accidentally, causing her pain from the memories of these lost loved ones and in relation to her own condition.

> *R: Sometimes I think about my parents, so I feel restless, that time it feels like I’m not able to take a breath, it feels like I’m not breathing, once I got senseless, I was very sick, my sister got to know about it, and she came and she took me to the hospital*
>
> *I: apart from your parent’s incident, is there anything else that makes you feel restless?*
>
> *R: About my husband, the death of my husband, I think about it a lot, as we stayed together, now he is no more, I can also die, first my parents died, I was living with my parents, they died, then I was living with my husband, but he also died, I can also die. (…) Nowadays I am always thinking about my after-death situation. (mother 12)*

### Isolated woman

Another woman, widow who work as maid and live in the tin house explained how she was surprised how the water level raised suddenly and how stressed she was by the situation.

> *R: Yes, I was afraid, suddenly waterlogged inside the house and also inside the road, there was no one here, only 3-4 people were here whose stuff was here, I wasn’t able to organize anything. (mother 13)*

The respondent was very tensed with exacerbated symptoms due to rumors in the slum about the occurrence of a new cyclone following Sitrang.

> *I: Still, you’re afraid because of this?*
>
> *R: Yes, people are saying that another cyclone is coming*
>
> *I: So, you are feeling afraid?*
>
> *R: Yes, I’m feeling afraid. I’m not going to work for 3-4 days. I can’t sleep, my body got wet, everything got wet. I got scared of seeing the water level, this is a problem! (mother 13)*

Another respondent also exhibits symptoms of anxiety and has difficulty sleeping at night due to the fear of new cyclones. This fear and anticipation of potential disasters can exacerbate mental health issues, making it difficult for individuals to focus on their daily tasks and responsibilities.

> *R: Sometimes at midnight, like in the night-time, I feel scared, like maybe suddenly it can come, because I’m staying in the tong house because nowadays people are saying a new one is coming, so that’s why I’m afraid if it happened again then what I’ll do. (mother 14).*

### Pathways of cumulative effects on mental health: a case study

The following case is that of a woman who represents a typical example of the cumulative effects of environmental, economic, and negative feedback loops when climate events occur on the mental health. The negative feedback loop typically intertwines with economic strategies related to the household size, such as migration and early marriage, or in some cases, deciding which children will migrate to the city or stay in the village, and eventually planning to return home.

##### Trajectory of a woman from the coastal area and feeling trapped in the slum

This woman was born in a coastal area of Bangladesh, exposed to risks associated with climate events such as cyclones, river erosion, and slow-onset events. She belonged to a family of farmers who owned multiple lands for rice crops and vegetables and had two houses. During her childhood, she witnessed numerous cyclones and storms. In the early 2000s, a cyclone destroyed one of her family’s houses, but no one was injured. She occasionally suffered from nightmares involving storms destroying everything, especially when her area was flooded and waterlogged. Since the road was reconstructed and reduced waterlogging, she no longer suffers from nightmares.

In the mid-2000s, her family fell into poverty after river erosion destroyed their lands and houses. With six children, her father decided, for economic reasons, to marry his two older daughters to men from the capital, hoping for better living conditions. Her older sister moved to Dhaka and married a man from the slum through her uncle’s intervention. Later, her sister brought her to Dhaka and married her to her husband.

After her marriage, she discovered that her husband was addicted to marijuana and alcohol, spending significant amounts of money on drugs. Her husband worked as a rickshaw driver, earning about 500-600 takas every day. However, he sold his rickshaw six days before our visit. The couple has many expenses, including education fees for their two children in primary school (300 takas every month). They live in a semi-built house in the slum near the lake, paying an expensive rent of around 4,000 takas for one room. Food expenses for the whole family amount to around 10-12,000 takas.

Her husband’s drug addiction created conflicts within their marriage, and he often physically abused her. Her mother-in-law, aware of the situation, protected her son, leaving the woman feeling isolated. The Cyclone Sitrang exacerbated her stress, and she expressed “feeling very stressed with tension all the time” about her children’s future.

This case shows how the woman’s trajectory has been influenced by climate events, her family’s economic problems, and the decisions made to cope with these challenges, such as migration and early marriages. Moreover, it highlights how persistent issues, such as poverty, addiction, and family conflicts, can worsen a woman’s mental health situation, especially when she feels trapped and isolated. This case also demonstrates that women are often the most affected by the consequences of climate events and economic decisions made within their households.

Other respondents mentioned after the cyclone Sitrang their sadness about leaving a child in the village as a survival strategy in order to move to the city, only to realize they had fallen into a dead-end trap. These climatic events reawaken past pains and impact mental health by highlighting how their strategies have ultimately turned against them.

The lack of opportunity impacts all members of these communities, but it appears to be especially pronounced among women who shoulder caregiving responsibility, have limited access to education, and occupy the lowest positions within family hierarchies. Both acute and slow-onset events distinctly affect the mental health of women, mothers, and widows, as it is influenced by the interplay of socioeconomic factors, social networks, and environmental conditions. These intersecting factors intensify the vulnerabilities these women experience, making it even more difficult for them to cope with the consequences of climate-related events.

## Discussion

Our study presents original findings on how women are disproportionately affected through two main mechanisms: socioeconomic stressors and acute environmental event. socioeconomic stressors, particularly during slow-onset environmental transformations, highlight the intersection between economic and environmental dimensions. Acute events directly cause psychological distress. In both cases, gender-related social status and roles shape individual trajectories. We specifically highlight that in these communities, economic survival strategies are based on the involvement of women in serving the household, which makes women’s health even more vulnerable during climate and environmental events. This exacerbates gender inequities and jeopardizes their mental health. Furthermore, our study allows for an analysis of cumulative environmental, economic, and social events by examining individual trajectories, opening up new perspectives for mental health research in the Global South. This approach allows us to identify both the structural sociocultural factors and the individual psychological factors that shape a person’s mental health trajectory.

The specificities of climate events are important to consider for the policy maker and in order to help the most vulnerable people. Usually the main characteristics retained are the duration, frequency, intensity of an event, geographical location such as coastal areas [31]. Our results showed the difference in perception and experiences lived by the most vulnerable for an event considered as of small-scale by the general authorities. While not of importance for the general population, it greatly affected both the living and economic conditions of a vulnerable community and their mental health. It is crucial for decision-makers to anticipate the impacts based on the topographic vulnerabilities of the sites and the individuals, in order to promote the establishment of shelters and the provision of material aid, even during less intense events such as Cyclone Sitrang. This situation is encountered by both men and women; however, it adds to gender-specific vulnerabilities [32] and should be taken into account to grasp the cumulative impact of vulnerabilities on individuals’ life trajectories.

Understanding the gender-specific vulnerabilities related to climate events is important to address the cumulative impact of vulnerabilities on individuals’ life trajectories. Mental health outcomes linked to SDGs will be difficult to achieve for women if the climate issue is not taken into account as it disproportionately affects women by reinforcing their vulnerabilities linked to social structures and producing new gender inequities. More specific studies should be made in the global souths because the indirect impacts of climate change on mental health are mostly uncovered as it happened mostly in the rural setting. Yet, these countries are the more vulnerable to climate events even if they are from far less responsible of the changing climate [33].

Our results highlight the importance for future research to consider psychological trajectories while taking into account health determinants, social factors, as well as environmental factors. This approach aims to better understand the mechanisms that affect populations and those shaping their resilience when they face several challenges. Our analysis confirms some other researcher based in Nigeria during floods in 2011. They found that this event impacted the childcare of women’s activities leading to increased anxiety about the health and safety of their children [34]. We also find in accordance with the literature various determinants that affect the link between mental health and climate change among women. Notably, one of the key factors is the socioeconomic status of women [35], which can exacerbate their vulnerability to the mental health impacts of climate change. Other determinants include gender roles and expectations, gendered violence [36], limited access to resources [37] and services after natural disaster [38], education, and the disproportionate burden of caregiving responsibilities [36]. These factors can contribute to increased stress, anxiety, psychological trauma and depression among women when faced with the challenges posed by climate change [12]. We demonstrate how the combination of factors such as socioeconomic status, gender roles, limited resources, and caregiving responsibilities can create a complex web of vulnerabilities that disproportionately affect women’s mental health in the face of climate change. By examining these interconnected factors in detail, we provide a more comprehensive understanding of the gendered impacts of climate change on mental health in vulnerable communities, thereby offering valuable insights for future research and policy interventions. Further research employing a trajectory-based approach is needed to better understand the interplay of factors and their contributions to the impacts of climate change on women’s mental health in vulnerable communities.

Our study has limits. Our interviews were conducted in Bengali and transcribed so we might have lost a part of the information. However, the analysis was performed with the translator to overcome this risk. Our data were collected on a two-period basis, while this topic is a sensitive one and might need iterative interviews to build a greater trust from the respondents. We tried to overcome this limit by triangulating within the same household and meeting multiple time with our respondents to build a trusting relationship.

We were perceived as foreigners and outsiders which might have had an impact on the quality of data. In both setting we have been associated to their survival economical strategies and the interviews could have been biased to meet the respondent needs. The tools from the reflexive ethnography have help us to analyze our position in each situation and how it has shaped the data.

## Conclusion

It is crucial to consider the specific mental health impacts of climate change on vulnerable groups, such as women, mothers, and widows, particularly in rural areas where resources and support may be limited. Providing access to mental health care, financial assistance, and climate-resilient livelihoods can help alleviate the strain on these individuals and improve their overall well-being.

Addressing the specific needs and vulnerabilities of women in the context of climate change requires a multi-faceted approach that includes promoting gender equality, empowering women to make informed decisions about their lives, and providing access to resources and support networks that can help them build resilience and cope with the challenges they face. Additionally, it is crucial to engage communities and stakeholders in the process of challenging and changing harmful gender norms and expectations that contribute to the vulnerability of women in the face of climate change.

## Data Availability

Due to the sensitive nature of the mental health data concerning women and in order to maintain anonymity and protect the privacy of the participants, the interview data cannot be shared.

## Acknowledgments

The authors acknowledge those who supported this work at various phases of our project and specifically all the participants on the two settings.

